# Redefining ischemic core, penumbra, and target mismatch on CT perfusion in acute anterior distal medium vessel occlusion

**DOI:** 10.1101/2025.03.25.25324574

**Authors:** Leon Y. Cai, Meisam Hoseinyazdi, Dhairya A. Lakhani, Hamza Salim, Janet Mei, Adam A. Dmytriw, Adrien Guenego, Thanh N. Nguyen, Shyam C. Majmundar, Richard Leigh, Elisabeth B. Marsh, Rafael H. Llinas, Victor C. Urrutia, Argye E. Hillis, Jens Fiehler, Gregory W. Albers, Jeremy J. Heit, Tobias D. Faizy, Vivek S. Yedavalli

## Abstract

**Background:** Recent trials of endovascular thrombectomy (EVT) for acute distal medium vessel occlusions (DMVOs) were negative but also used inconsistent imaging-based inclusion criteria, whereas many successful large vessel occlusion (LVO) EVT trials used empirically validated CT perfusion-based target mismatch (TMM) criteria: an ischemic penumbra (time-to-maximum [Tmax] >6s) to core (relative cerebral blood flow [rCBF] <30%) mismatch ratio (MMR) ≥1.8 and volume (MMV) ≥15mL. We aimed to determine optimal corresponding definitions in DMVOs to improve patient selection for EVT.

**Methods:** We retrospectively analyzed patients with acute anterior DMVOs from prospectively collected databases at four comprehensive stroke centers. To assess core, we evaluated how well pretreatment rCBF <20%, <30%, <34%, and <38% volumes correlated with MRI-based post-treatment follow-up volumes (FIVs) in successfully recanalized patients. To evaluate penumbra, we assessed how well pretreatment Tmax >4s, >6s, >8s, and >10s volumes correlated with FIVs in unrecanalized patients. Then, we evaluated whether these improved parameters for core and penumbra better quantified LVO TMM and identified an optimal DMVO TMM definition.

**Results:** In 122 core patients, rCBF <38% most strongly correlated with FIVs (concordance correlation coefficient [CCC] 0.30 [95% CI, 0.15-0.48]), outperforming rCBF <30% (CCC 0.21 [0.10-0.35]) (p < 0.001). In 70 penumbra patients, Tmax >8s most strongly correlated with FIVs (CCC 0.49 [0.25-0.77]), outperforming Tmax >6s (CCC 0.39 [0.17-0.68]) (p < 0.001). In 180 patients undergoing EVT with Tmax >6s to rCBF <30% MMR ≥1.8 and MMV ≥15mL, recomputing MMR and MMV using Tmax >8s and rCBF <38% further separated those with favorable outcomes (p = 0.007). In the same cohort, Tmax >8s to rCBF <38% MMR ≥2.2 and MMV ≥10mL maximized the number of patients benefitted (p < 0.001, absolute risk reduction 26%).

**Conclusions:** In acute anterior DMVOs, rCBF <38% and Tmax >8s best correspond to ischemic core and penumbra, respectively; more favorably quantify LVO TMM; and reveal optimal TMM criteria. These results should be prospectively investigated as inclusion criteria for EVT in this population and suggest recent negative DMVO EVT trials may have been confounded by suboptimal patient selection.

## Introduction

Following multiple randomized controlled trials, endovascular thrombectomy (EVT) has emerged as the standard of care for patients with acute large vessel occlusion (LVO) ischemic stroke for up to 24 hours after symptom onset^1–7^. Previous studies have found that patients who present with a “ target mismatch” (TMM) on pretreatment perfusion imaging, typically with CT perfusion (CTP) or MRI, are most likely to benefit from EVT. TMM has been defined as a joint ischemic perfusion deficit volume to ischemic core volume ratio (i.e., “ mismatch ratio” or MMR) of ≥1.8 and volume difference (i.e., “ mismatch volume” or MMV) of ≥15mL^6,8,9^. These thresholds are thought to characterize the ischemic penumbra, or remaining salvageable tissue with a perfusion deficit that has not yet progressed to ischemic core. Of note, the term “ penumbra” is often used to describe the perfusion deficit itself, and we will refer to it as such herein.

In part due to the unquestionable benefit of EVT for LVO, studies have evaluated EVT for acute distal medium vessel occlusion (DMVO) ischemic strokes as well, with early and small studies finding mixed to some benefit^10–12^. Recent large clinical trials, however, have found neutral to negative outcomes with EVT in DMVO^13,14^. In addition to combining anterior and posterior occlusions, one factor that in part may be responsible for these conflicting results is that the trials largely lacked consensus regarding the perfusion imaging criteria for EVT triage^15^. For instance, one trial defined penumbra as the time-to-maximum (Tmax) >4s volume and core as hypodensity on CT without contrast, setting mismatch criteria at a core-to-penumbra ratio of <0.9 (defined reciprocally to MMR)^14,15^. Another trial also used Tmax >4s for penumbra, but defined core as the relative cerebral blood flow (rCBF) <30% volume with mismatch criteria set at a core-to-penumbra ratio of <0.5 (Rapid Medical, Yokneam, Israel)^15^. A third trial left the protocol definition of “ mismatch” open, and a fourth did not use perfusion-based criteria at all^13,15,16^. Notably, these criteria have not been validated for DMVOs and were all different than the rCBF <30% volume definition for core and Tmax >6s definition for penumbra with TMM MMR ≥1.8 and MMV ≥15mL that has been repeatedly validated as inclusion criteria for EVT in LVOs^6,7,17,18^.

Thus, despite the negative trial results for EVT in DMVOs, there remains a question as to whether outcomes would have been different with stricter perfusion imaging-based patient selection criteria. To answer this question, one logical step would be to utilize the validated LVO definitions of ischemic core, penumbra, and TMM in a trial for DMVOs^6,8,9,19,20^. However, prior retrospective studies have suggested that rCBF <30% and Tmax >6s may be suboptimal parameters for measuring core and penumbra, respectively, in DMVOs; and there is minimal literature regarding whether the LVO TMM definition is optimized for DMVOs^21^. Further, additional studies have suggested patient selection for EVT in this population must be more stringent to avoid complications^22^. Therefore, we posit that without a full understanding of how ischemic core, penumbra, and TMM are reflected in perfusion imaging in patients with DMVOs, our ability to optimally design imaging-based inclusion criteria for future trials in this population is limited.

As such, in this work we aim to evaluate not only which CTP parameters best estimate ischemic core and penumbra in acute anterior DMVOs, but also aim to understand which CTP TMM patient selection criteria are best for reperfusion therapy in this population. Specifically, we seek to understand which rCBF and Tmax volumes yield the best estimations of core and penumbra volumes, respectively, in DMVOs and how those estimates compare to the rCBF <30% and Tmax >6s volumes traditionally used for LVOs. Additionally, we seek to investigate which rCBF and Tmax volumes better stratify outcomes under traditional LVO-defined TMM inclusion criteria in DMVOs as well as whether there exists a more optimal TMM definition for acute anterior DMVOs altogether.

## Methods

### Study design

For this study, we retrospectively analyzed a database of patients seen at four comprehensive stroke centers between 2010 and 2024: Johns Hopkins Hospital and Johns Hopkins Bayview Medical Center (Baltimore, MD, USA), Stanford University Medical Center (Palo Alto, CA, USA), and University Medical Center Hamburg-Eppendorf (Hamburg, Germany). These data were originally prospectively collected and continuously maintained in databases. Patients were included following the respective Institutional Review Board guidelines, regulations, and requirements (Hopkins 00269637, Stanford 37209, and Hamburg 689-15). This study was conducted according to the Strengthening the Reporting of Observational Studies in Epidemiology (STROBE) guidelines. The data supporting this study are available from the corresponding author upon reasonable request.

### Inclusion and exclusion criteria

The inclusion criteria varied for different portions of this study, and three overall groups were identified. Criteria common to all were (1) successful pretreatment CT without contrast, CT angiography (CTA), and CTP without significant artifact from motion, inadequate contrast timing, or other technical failure; and (2) primary anterior circulation DMVO on pretreatment CTA defined as an M2-, M3-, or M4-segment middle cerebral artery occlusion or anterior cerebral artery (ACA) occlusion as determined by an experienced neuroradiologist (VSY, 10 years of experience). Patients with tandem occlusions or concomitant LVOs were excluded.

Of the three groups, the first two were a “ core” and “ penumbra” group for volume estimation, respectively. For the core group, we sought to identify patients who were successfully recanalized, indicating their pretreatment estimations of core on CT perfusion would be most theoretically equivalent to follow-up infarct volumes (FIVs) measured on post-treatment MRI. Thus, criteria for the core group also included successful recanalization with either intravenous thrombolysis (IVT) or EVT with achievement of a modified thrombolysis in cerebral infarction (mTICI) score ≥2b on intraoperative digital subtraction angiography (DSA) or follow-up CTA or MR angiography (MRA) in cases of reperfusion with IVT alone.

For the penumbra group, we sought to identify patients who were not successfully recanalized, indicating their pretreatment estimations of penumbra on CT perfusion would be most equivalent to FIV on post-treatment MRI. Thus, criteria for the penumbra group included those patients who did not undergo reperfusion therapy or underwent IVT or EVT but without successful recanalization, defined as mTICI ≤2a on intraoperative DSA.

As such, additional common criteria to these two groups were (1) successful post-treatment MRI, specifically diffusion weighted imaging (DWI) or fluid attenuated inversion recovery (FLAIR) sequences, without significant artifact from motion or other technical failure; and (2) monotonically changing volumes at the different CTP parameter thresholds defined below.

The third group was a “ TMM analysis” group of DMVO patients who underwent EVT using the standard-of-care LVO TMM definition (i.e., Tmax >6s to rCBF <30% MMR ≥1.8 and MMV ≥15mL). This group was used to characterize how new core and penumbra measurements and TMM definitions compared to the existing LVO measurements and definition and to assess which may be optimal in patients with DMVO undergoing EVT. Inclusion criteria for this group required (1) patients undergo EVT regardless of mTICI outcome and (2) have a valid 90-day modified Rankin Scale (mRS) or discharge National Institutes of Health Stroke Scale (NIHSS) as measured by a board-certified stroke neurologist or nurse practitioner.

### Imaging data

Pretreatment CTP parameters investigated included the volume of rCBF <20%, <30%, <34%, and <38% as well as volume of Tmax >4s, >6s, >8s, and >10s. All volumes were automatically computed from CTP with RAPID AI software (iSchemaView, Menlo Park, CA, USA) and subsequently quality checked with artifactual volumes corrected or removed.

Post-treatment FIVs were semi-automatically computed from DWI when available and FLAIR otherwise within 48 hours of EVT for successfully recanalized patients or 72 hours of symptom onset for unrecanalized patients. In cases where MRI was not performed, follow up CT without contrast was used in the same timeframe. Segmentation was performed by two board-certified neuroradiologists (VSY, 10 years of experience; MH, 4 years of experience) using the Phillips Carestream Vue PACS (Cambridge, MA, USA) segmentation tool. FIVs were manually quality checked with artifactual volumes corrected or removed.

All volumes are reported in mL.

### Clinical data

Each patient’s age, sex, admission NIHSS, Alberta Stroke Program Early CT Score (ASPECTS), and time from last known well (LKW) to pretreatment CTP were also recorded as was whether the patient received IVT or EVT. Additionally, each patient’s medical history regarding smoking, hypertension, hyperlipidemia, diabetes, coronary disease, atrial fibrillation, and use of antiplatelet or anticoagulants were also recorded as were the presenting systolic blood pressure, blood glucose, and platelet count.

### Outcome definitions

Our primary outcome measure for the core and penumbra estimations was FIV. The primary outcome measure for TMM characterization was a composite clinical outcome, with favorable defined as a 90-day mRS of 0-2 or a discharge NIHSS of 0-1^8^.

### Statistical analysis

Univariate differences between groups were analyzed as indicated. For continuous variables, statistically significant differences were computed with the Wilcoxon rank-sum test. For categorical variables, statistically significant differences were determined with Pearson’s chi-squared test.

To qualitatively assess agreement between pretreatment volume estimates and FIVs, Bland-Altman plots were used. To quantitatively assess agreement, unadjusted coefficients of determination (R^2^) and concordance correlation coefficients (CCCs) were computed. Distributions for the CCCs were computed with bootstrapping with 1000 iterations and compared between different pretreatment volume estimates with one-way analyses of variance (ANOVAs) and subsequent post-hoc Tukey’s tests. Linear slope-intercept models relating pretreatment volume estimates to FIV were also computed.

Throughout, statistical significance was set at p < 0.05 without multiple comparisons correction.

### Optimization analysis

To identify the search range for an optimal TMM definition for DMVOs, percent favorable outcome in patients included across candidate MMRs and MMVs (i.e., patients with MMR or MMV greater than or equal to a given candidate MMR or MMV, respectively) was plotted. The search ranges for the optimal MMR and MMV were each then identified as the range of candidates that produced a qualitatively higher and increasing percentage of favorable outcomes than the reference LVO definition. This search range for the MMR and MMV were then used jointly to optimize the TMM definition by identifying the MMR and MMV combination that maximized the number of patients benefitted (Eq. 1):

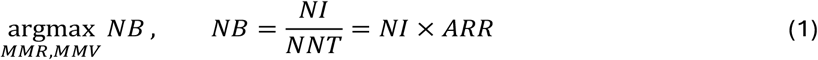

where *NB* is the number of patients benefitted, *NI* is the number of patients included by the joint criteria, *NNT* is the number needed to treat (equivalent to the number needed to benefit), and *ARR* is the absolute risk reduction in unfavorable outcomes between those who meet the joint criteria and those who do not (equivalent to the absolute percent increase in benefit between those groups).

## Results

A total of 1593 patients were screened for this study with 122 included in the core estimation group (120 M2 and 2 M3), 70 in the penumbra estimation group (63 M2, 4 M3, 1 M4, and 2 ACA), and 180 in the TMM analysis group (175 M2 and 5 M3) (Table 1).

**Table 1.** Baseline data for patients included in the core and penumbra estimation groups with statistical comparison as well as an overall summary for the TMM analysis group.

|  | Penumbra<br>N=70 | Core<br>N=122 | p | TMM<br>N=180 |
| --- | --- | --- | --- | --- |
| **Occlusion Site** |  |  |  |  |
| M2 | 63 | 120 | — | 175 |
| M3 | 4 | 2 | — | 5 |
| M4 | 1 | 0 | — | 0 |
| ACA | 2 | 0 | — | 0 |
| **Clinical Data** |  |  |  |  |
| Age | 70.0 (19.0) | 75.0 (17.0) | 0.109 | 76.0 (17.0) |
| Sex (male) | 31 (44.3) | 69 (56.6) | 0.101 | 88 (48.9) |
| NIHSS | 9.0 (13.0) | 10.0 (11.0) | 0.333 | 10.5 (11.0) |
| LKW (hours) | 6.3 (9.7) | 2.2 (4.6) | <0.001 | 3.1 (5.6) |
| ASPECTS | 8.0 (3.0) | 9.0 (3.0) | 0.162 | 9.0 (2.0) |
| Systolic blood pressure | 154.0 (43.2) | 153.0 (34.5) | 0.938 | 153.0 (40.0) |
| Glucose | 121.0 (36.2) | 121.5 (49.0) | 0.877 | 122.0 (40.8) |
| Platelet count | 216.0 (85.0) | 200.0 (122.5) | 0.413 | 202.0 (118.2) |
| **Medical History** |  |  |  |  |
| Smoking | 35 (50.7) | 37 (31.9) | 0.011 | 53 (30.6) |
| Hypertension | 55 (78.6) | 97 (79.5) | 0.878 | 143 (79.4) |
| Hyperlipidemia | 26 (40.0) | 45 (40.5) | 0.944 | 71 (43.8) |
| Diabetes | 13 (18.6) | 32 (26.2) | 0.228 | 44 (24.4) |
| Coronary disease | 21 (41.2) | 34 (46.6) | 0.552 | 58 (53.2) |
| Atrial fibrillation | 18 (25.7) | 55 (45.8) | 0.006 | 78 (43.8) |
| Anti-platelet or -coagulant | 26 (37.1) | 56 (46.3) | 0.219 | 80 (44.7) |
| **Interventions** |  |  |  |  |
| IVT | 12 (17.1) | 63 (52.1) | <0.001 | 83 (46.4) |
| EVT | 30 (42.9) | 122 (100.0) | <0.001 | 180 (100.0) |
| **Outcomes** |  |  |  |  |
| 90-day mRS 0-2 | 26 (37.1) | 59 (48.4) | 0.132 | 78 (43.3) |
| Discharge NIHSS 0-1 | 13 (18.6) | 39 (32.0) | 0.044 | 45 (25.0) |
| Composite | 29 (41.4) | 66 (54.1) | 0.091 | 87 (48.3) |

### Core and penumbra volume analysis

Baseline differences between the core and penumbra groups are reported in Table 1. Notable differences include a median time since LKW of 6.3 hours in the penumbra group and 2.2 in the core group (p < 0.001), more smokers (50.7%) in the penumbra group compared to 31.9% in the core group (p = 0.011), and more atrial fibrillation in the core group (45.8%) compared to 25.7% in the penumbra group (p = 0.006) in the core group. As expected, more patients received IVT at 52.1% and EVT at 100.0% in the core group compared to the penumbra group at 17.1% and 42.9%, respectively (p < 0.001 for both). Similarly, more patients in the core group (32.0%) had a discharge NIHSS of 0-1 compared to the penumbra group (18.6%) (p = 0.044). Importantly, no statistical differences in age or admission NIHSS were noted.

The Bland-Altman plots relating pretreatment CTP estimates to FIV for both the core and penumbra groups are provided in Figure 1. For the core estimates, increasing rCBF threshold demonstrated mean differences closer to zero, improving from −30.76 at <20%, to −22.49 at <30%, to −17.84 at <34%, and to −13.13 at <38%. The variance remained similar across all thresholds with 1.96 times the standard deviation (1.96σ) sitting between 90 and 95.

**Figure 1.**
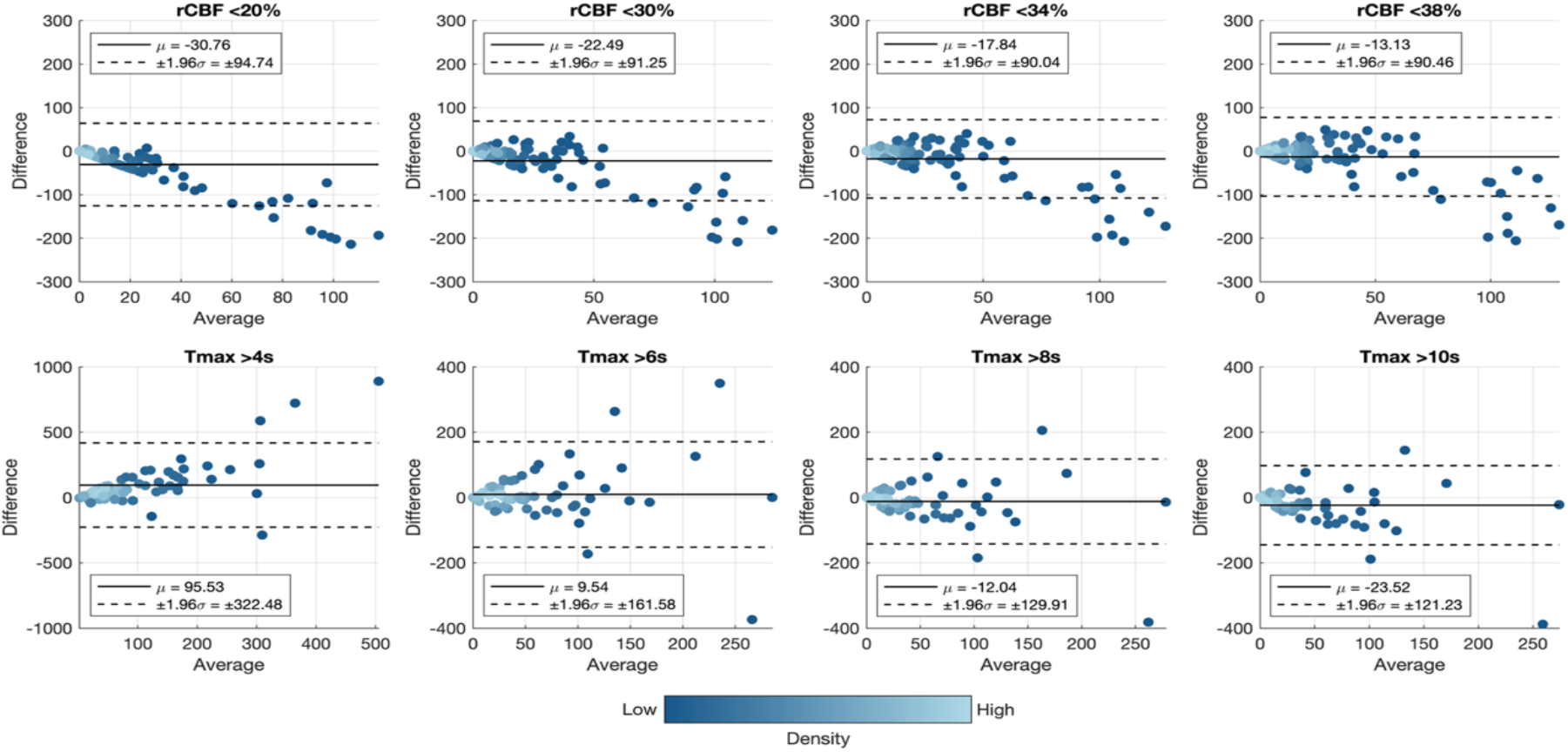
Qualitative analysis of agreement between pretreatment CTP parameters and FIV with Bland-Altman plots in the core (rCBF) and penumbra (Tmax) groups.

For the penumbra estimates, Tmax >6s had the lowest absolute mean difference at 9.54. Tmax >8s had a comparable mean difference at −12.04 with notably lower variance, with a 1.96σ of 129.91 down from 161.58 for Tmax >6s. Tmax >10s had even lower variance with 1.96σ of 121.23 but a mean difference with increased negative bias at −23.52. Tmax >4s had dramatically larger variance and mean difference compared to all others.

Quantitatively, the R^2^ for the core and penumbra estimates compared to FIV were computed. For core, rCBF <38% had the highest R^2^ at 0.10, followed by 0.06 for rCBF <34%, −0.04 for rCBF <30%, and −0.28 for rCBF <20%. For penumbra, Tmax >10s had the highest R^2^ at 0.18, followed closely by Tmax >8s at 0.16. Tmax >6s had an R^2^ of −0.28, and Tmax >4s had an R^2^ of −5.75.

Scatter plots relating pretreatment CTP estimates to FIV are provided in Figure 2. For the core estimates, increasing rCBF threshold was associated with increasing CCC, improving from 0.09 at <20%, to 0.21 at <30%, to 0.26 at <34%, and to 0.30 at <38%. These increases were statistically significant (Figure 3). The lines of best fit also visually agreed with these findings as they approached y=x with increasing rCBF threshold (Figure 2).

**Figure 2.**
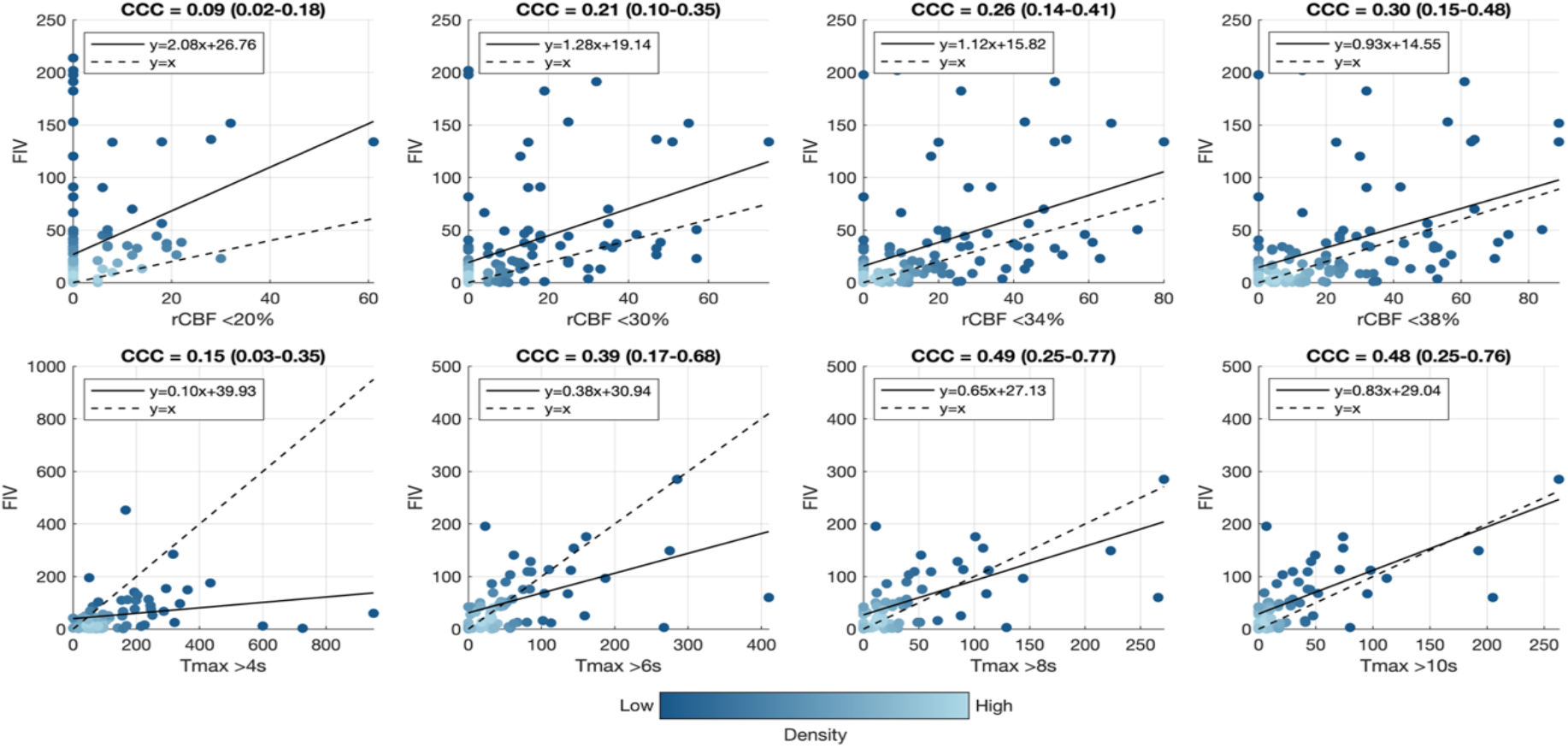
Quantitative analysis of agreement between pretreatment CTP parameters and FIVs in the core (rCBF) and penumbra (Tmax) groups.

**Figure 3.**
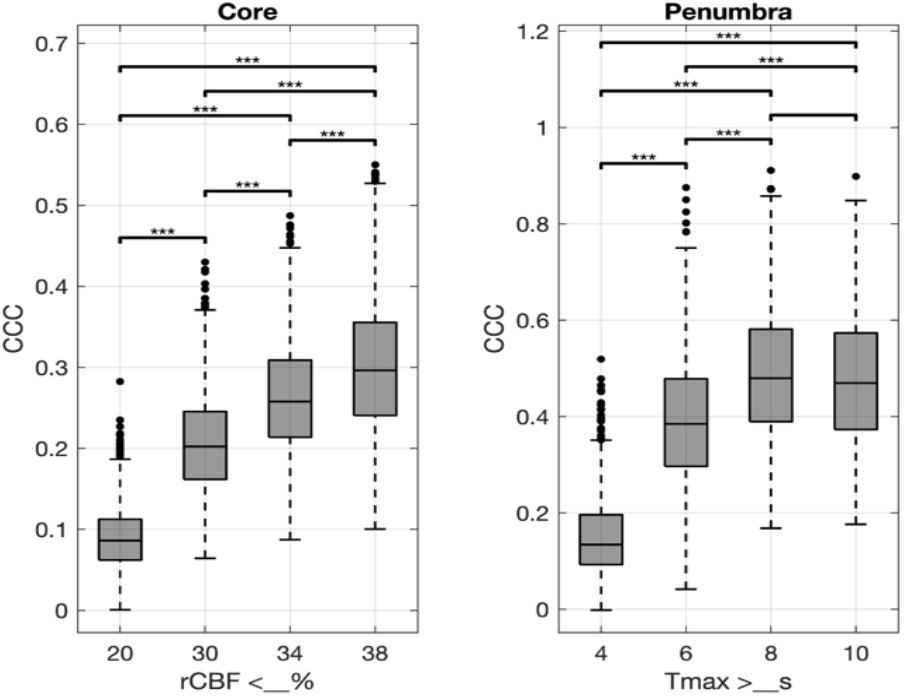
Statistical comparisons of CCC between different pretreatment CTP parameters and FIVs for core and penumbra estimation. *** p < 0.001.

For the penumbra estimates, Tmax >8s achieved the highest CCC at 0.49, followed by Tmax >10s with 0.48, then Tmax >6s with 0.39, and finally Tmax >4s with 0.15 (Figure 2). These decreases were statistically significant with the exception of the Tmax >8s and >10s comparison (Figure 3). The lines of best fit visually agree with these findings as those of Tmax >8s and Tmax >10s most closely approximate y=x (Figure 2).

### Target mismatch definition analysis

Baseline characteristics for the TMM analysis group as a whole are reported in Table 1. Notably, the median age was 76, the median admission NIHSS was 10.5, the median time since LKW was 3.1 hours, and 46.4% of patients also received IVT. All patients received EVT. The overall percentage of favorable outcomes was 48.3% (Table 2). This percentage represents a reference for DMVO patients who underwent EVT using traditional LVO TMM criteria with rCBF <30% and Tmax >6s volumes and MMR ≥1.8 and MMV ≥15.

**Table 2.** Favorable outcomes by core-penumbra measurement and TMM definition.

| Measurement |  | TMM Definition |  | Included | Favorable Outcomes |  |  |
| --- | --- | --- | --- | --- | --- | --- | --- |
| Core | Penumbra | MMR | MMV | N (% of total) | N (% of included) | N (% of excluded) | p |
| rCBF <30% | $T_{max} >6s$ | $\geq 1.8$ | $\geq 15$ | 180 (100.0) | 87 (48.3) | N/A (Reference) | N/A |
| rCBF <38% | $T_{max} >8s$ | $\geq 1.8$ | $\geq 15$ | 93 (51.7) | 54 (58.1) | 33 (37.9) | 0.007 |
| rCBF <38% | $T_{max} >8s$ | $\geq 2.2$ | $\geq 10$ | 90 (50.0) | 55 (61.1) | 32 (35.6) | <0.001 |

Estimating ischemic core with rCBF <38% and penumbra with Tmax >8s and under a traditional TMM definition of MMR ≥1.8 and MMV ≥15, 93 of the 180 patients in the group (51.7%) met criteria (Table 2). Of these 93 patients, 54 (58.1%) had a favorable outcome compared to 33 of the 87 (37.9%) who did not meet criteria (p = 0.007). This effect in composite outcome was paralleled by a higher percentage of included patients with a 90-day mRS of 0-2 at 50.5% compared to 35.6% in the excluded group, but no differences between rates of NIHSS 0-1 at discharge were found (Supplemental Table S1).

Between the included and excluded patients, 60.2% of those included were men compared to 36.8% in the excluded group (p = 0.002), and 54.3% of patients included had atrial fibrillation compared to 32.6% in the excluded group (p = 0.003). No other statistically significant differences in covariates were noted (Supplemental Table S1).

The optimization process to identify the optimal TMM definition using rCBF <38% and Tmax >8s is provided in Figure 4. The search range for MMR was visually determined to be 1.2 to 2.6, as these ratios found increasingly favorable outcomes over the reference prior to a peak effect at 2.6 and plateau-to-decreasing effect when >2.6 (Figure 4a, left). Similarly, the search range for MMV was visually determined to be 5 to 42, as these volumes found increasingly favorable outcomes over the reference prior to a peak effect observed at a volume of 42 (Figure 4a, right)

**Figure 4.**
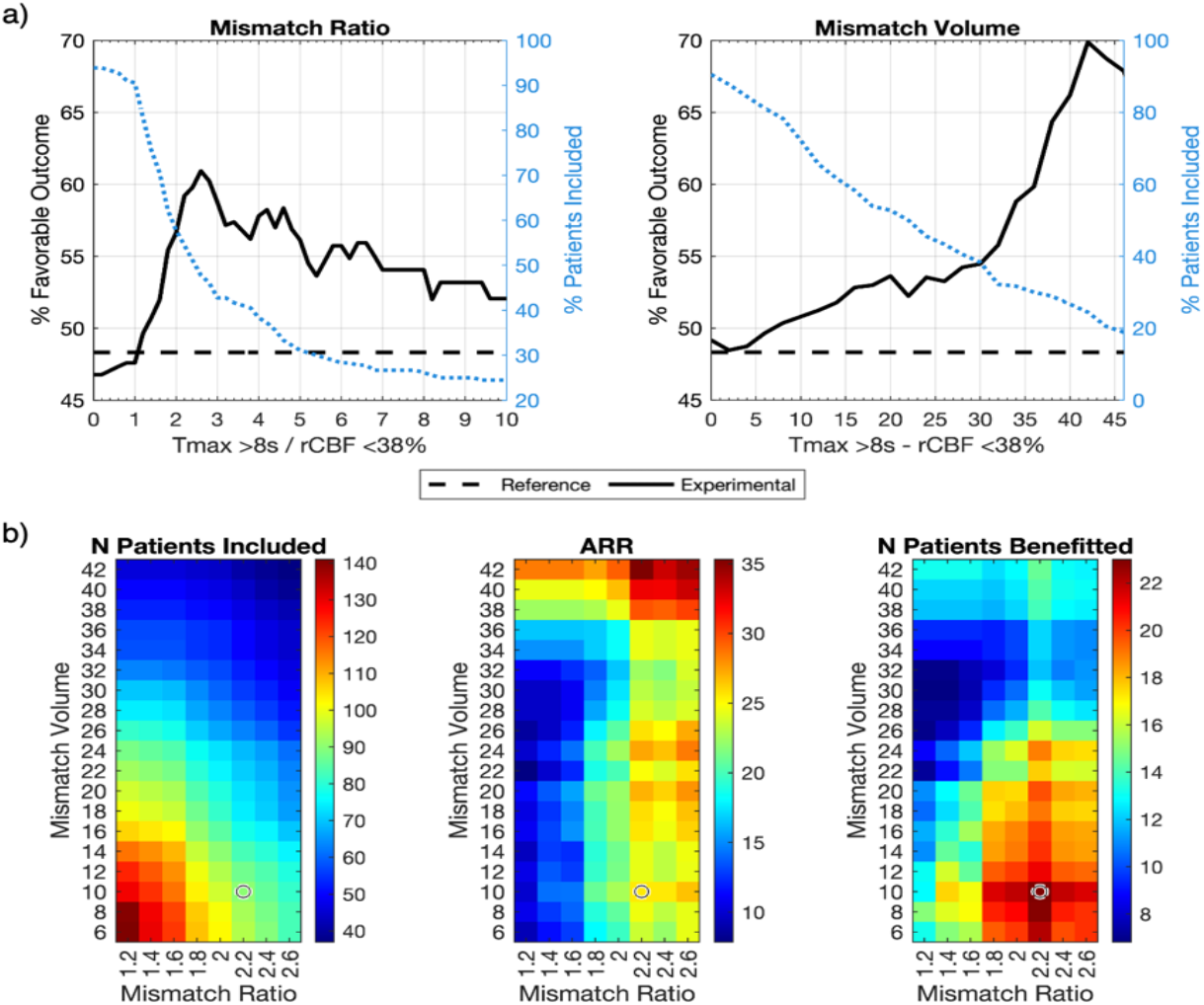
Identifying an optimal TMM definition for acute DMVOs. (a) Percent favorable outcomes in patients with a Tmax >8s to rCBF <38% MMR and MMV greater than or equal to candidate thresholds. The percent of patients included at each candidate threshold is also plotted. (b) Visualization of the optimization process with search ranges derived from the upslopes in (a). The optimal MMR and MMV thresholds (circled) maximize the number of patients benefitted, computed as the product of the number of patients included and the ARR (%).

Using these search ranges, the number of patients included by the joint criteria as a function of the MMR and MMV cutoffs were plotted (Figure 4b, left). The highest number of included patients was seen with low MMR and MMV cutoffs. The ARR was then also plotted, with highest ARR seen with higher MMR and MMV cutoffs (Figure 4b, center). The optimization function demonstrated the highest number of patients benefitted with high MMR and low MMV cutoffs, with the optimal criteria at an MMR of 2.2 and MMV of 10 (Figure 4b, right).

Under new TMM criteria of Tmax >8s to rCBF <38% MMR ≥2.2 and MMV ≥10, 90 (50.0%) patients were included (Table 2). Of these 90 patients, 55 (61.1%) had a favorable outcome compared to 32 (35.6%) of the 90 excluded (p < 0.001). This effect was consistent when broken down by outcome measure (Supplemental Table S1). Specifically, 53.3% of those included had a 90-day mRS of 0-2 compared to 33.3% in the excluded group (p = 0.007), and 33.3% of those included had a discharge NIHSS of 0-1 compared to 16.7% of those excluded (p = 0.010).

Between the included and excluded patients under these criteria, more men were in the included group (56.7%) than excluded group (41.1%) (p = 0.037), the included patients had a lower median NIHSS of 9 compared to 12 in the excluded group (p = 0.010), and the included patients had a higher percentage of patients with atrial fibrillation at 55.1% compared to 32.6% in the excluded group (p = 0.003). No other statistically significant differences in covariates were noted (Supplemental Table S1).

Regarding favorable outcomes in this cohort of patients with DMVO who underwent EVT, Tmax >8s to rCBF <38% MMR ≥2.2 and MMV ≥10 captured a sensitivity of 0.63, specificity of 0.62, positive predictive value of 0.61, negative predictive value of 0.64, and ARR of 26% with a NNT of 4.

## Discussion

In this multicenter study of acute anterior DMVOs, we first found in successfully recanalized patients that rCBF <38% volumes qualitatively exhibited the lowest bias in error against FIVs as well as the highest CCC and R^2^ compared to the other rCBF cutoffs. Second, we found in unsuccessfuly recanalized patients that Tmax >8s and Tmax >10s had similar CCC and R^2^ against FIVs, but Tmax >8s exhibited the lowest combination of bias and variance in error. Notably, both rCBF <38% and Tmax >8s saw improvement over rCBF <30% and Tmax >6s, respectively. Third, we found that in patients who underwent EVT under rCBF <30% and Tmax >6s MMR ≥1.8 and MMV ≥15mL, recomputing MMR and MMV with rCBF <38% and Tmax >8s further stratified favorable from unfavorable outcomes with minimal confounding. Finally, in this same cohort, we found that using rCBF <38% and Tmax >8s with MMR ≥2.2 and MMV ≥10mL maximized the number of patients benefitted by including half of patients with a strongly significant ARR of 26% and NNT of 4. Notably, the patients included by these new criteria had a lower median admission NIHSS than the excluded patients, but both fell within range of moderate severity strokes.

Thus, we conclude that rCBF <38% and Tmax >8s volumes best estimate ischemic core and penumbra in this population, respectively, as well as MMR and MMV, particularly when compared to rCBF <30% and Tmax >6s. We also conclude that a TMM definition of MMR ≥2.2 and MMV ≥10mL is most optimal for acute anterior DMVOs.

Compared to major LVO trials of EVT, perfusion imaging-based criteria in DMVO EVT trials were inconsistently utilized^6,7,17,18^. For instance, some of the DMVO trials utilized a penumbra definition of Tmax >4s but at the same time used different core definitions, either derived from CT without contrast or rCBF <30%^14,15^. Another left the definition of mismatch open^13^. Notably, we found that Tmax >4s is a poor estimation of penumbra and that similarly rCBF <30% is not optimal. Specifically, we found that, compared to rCBF <38%, lower rCBF thresholds had larger negative biases for core estimation and that compared to Tmax >8s, lower Tmax thresholds had larger positive biases. Thus, MMR and MMV computed with rCBF <20%, <30%, and <34% in conjunction with Tmax >4s and >6s in this population would likely be overestimated. As TMM is defined as a lower bound MMR and MMV, this would necessarily result in over-inclusion of patients for EVT, thus potentially biasing toward negative results. This is supported herein by further separation of favorable and unfavorable outcomes using rCBF <38% and Tmax >8s but the same LVO TMM definition. Thus, at the very least, the results herein highlight the need for consistent and optimized mismatch definitions between trials to reduce the impact of imaging-based patient selection criteria on outcomes. Specifically, we suggest that the proposed definitions of ischemic core, penumbra, and TMM (1) have promise as optimized parameters for acute anterior DMVO, (2) are superior to the LVO criteria in this population, and (3) should be evaluated in future prospective studies as inclusion criteria for EVT for these patients.

We note that the selection of inclusion criteria parameters often requires consideration beyond numerical “ optimality”. For instance, Cereda et al. found that rCBF <38% demonstrated the lowest mean absolute difference between CTP and FIV for core estimation in LVO, and the “ optimal” TMM MMR cutoff was original computed by Kakuda et al. as 2.6^8,19^. However, rCBF <30% and MMR ≥1.8 were ultimately chosen to avoid excluding patients with LVO from EVT^8,19^. Similarly, we found here that selecting a lower MMR cutoff for DMVO (Figure 4b, left) would also increase the number of patients included. However, the benefits of including more patients must be weighed against the reduction in favorable effect, as demonstrated by a steep drop in ARR (Figure 4b, center) with lower MMR. As such, the optimization criterion herein was set as the number of patients benefitted to balance these needs (Eq. 1).

One key takeaway from these results is that optimal parameters for DMVO prognosis and treatment efficacy may not be the same as those for LVOs. This is consistent with prior work suggesting collaterals may play a different or even outsized role in DMVOs comparatively^23–25^. It is also consistent with prior work indicating that imaging and clinical parameters unique to DMVOs ought to be investigated further as both prognostic and selective criteria^24,26–30^.

Notably, this study has limitations. This work is inherently limited by its retrospective design. The largest bias that arises as a result is regarding the TMM analysis group. By definition, the standard of care for these patients was to undergo EVT by LVO TMM criteria measured with rCBF <30% and Tmax >6s. Thus, all patients in this group meet these criteria by definition. This preempts an analysis of those who do not meet LVO TMM criteria but are taken for EVT under the proposed criteria, and thus we cannot do a full head-to-head comparison of the proposed criteria against established LVO criteria and can only use the latter as a reference. This limitation highlights the need to evaluate these thresholds prospectively.

An additional limitation is that the time between pretreatment CTP and post-treatment MRI was not constrained to be immediate (i.e, within hours). Thus, the negative bias between all rCBF estimates and follow-up FIVs in the core group likely reflects infarct growth in the interim which ought to be accounted for in future prospective studies. This time dependence is consistent with prior work^31^.

Further, the bulk of the patients included in this study had M2-segment occlusions. This is consistent with recent clinical trials where the plurality and majority of anterior circulation DMVOs identified were M2-segment occlusions^13,14^. However, compared to those trials, our study still includes a smaller proportion of non-M2-segment occlusions.

Overall, our study provides evidence that anterior DMVO-specific definitions of core and penumbra as well as TMM on pretreatment CTP show promise for improving triaging for reperfusion therapy for these patients. Additionally, our results suggest that optimization of ischemic core and penumbra parameters is worthwhile when designing future DMVO trials.

## Data Availability

The data supporting this study are available from the corresponding author upon reasonable request.

## Acknowledgements

None

