## Supplemental for "Redefining ischemic core, penumbra, and target mismatch on CT perfusion in acute anterior distal medium vessel occlusion"

**Supplemental Information**

**Supplemental Table S1.** Baseline data for patients included and excluded by different MMR and MMV thresholds computed using rCBF <38% and Tmax >8s volumes in the TMM analysis group.

| | MMR ≥1.8 and MMV ≥15mL | | | MMR ≥2.2 and MMV ≥10mL | | |
|---|---|---|---|---|---|---|
| | Excluded | Included | p | Excluded | Included | p |
| **Clinical Data** | | | | | | |
| Age | 75.0 (17.5) | 77.0 (16.0) | 0.586 | 75.0 (19.0) | 77.0 (15.0) | 0.347 |
| Sex (male) | 32 (36.8) | 56 (60.2) | 0.002 | 37 (41.1) | 51 (56.7) | 0.037 |
| NIHSS | 10.0 (11.0) | 11.0 (9.2) | 0.728 | 12.0 (12.0) | 9.0 (9.0) | 0.010 |
| LKW (hours) | 3.2 (5.6) | 2.9 (5.7) | 0.737 | 2.8 (5.5) | 3.3 (6.4) | 0.311 |
| ASPECTS | 9.0 (2.8) | 9.0 (2.0) | 0.267 | 9.0 (3.0) | 9.0 (2.0) | 0.316 |
| Systolic blood pressure | 153.5 (35.5) | 153.0 (44.0) | 0.956 | 153.0 (41.8) | 153.0 (38.2) | 0.978 |
| Glucose | 117.5 (44.0) | 122.0 (37.5) | 0.721 | 119.0 (41.8) | 122.0 (40.0) | 0.847 |
| Platelet count | 211.0 (119.8) | 202.0 (100.0) | 0.837 | 213.5 (118.0) | 199.0 (84.8) | 0.404 |
| **Medical History** | | | | | | |
| Smoking | 26 (31.7) | 27 (29.7) | 0.772 | 28 (32.9) | 25 (28.4) | 0.518 |
| Hypertension | 69 (79.3) | 74 (79.6) | 0.966 | 71 (78.9) | 72 (80.0) | 0.854 |
| Hyperlipidemia | 28 (37.8) | 43 (48.9) | 0.159 | 32 (41.6) | 39 (45.9) | 0.580 |
| Diabetes | 18 (20.7) | 26 (28.0) | 0.257 | 20 (22.2) | 24 (26.7) | 0.488 |
| Coronary disease | 25 (54.3) | 33 (52.4) | 0.839 | 30 (58.8) | 28 (48.3) | 0.271 |
| Atrial fibrillation | 28 (32.6) | 50 (54.3) | 0.003 | 29 (32.6) | 49 (55.1) | 0.003 |
| Anti-platelet or -coagulant | 36 (41.4) | 44 (47.8) | 0.386 | 38 (42.2) | 42 (47.2) | 0.504 |
| **Interventions (All received EVT)** | | | | | | |
| IVT | 39 (45.3) | 44 (47.3) | 0.792 | 40 (44.4) | 43 (48.3) | 0.604 |
| **Outcomes** | | | | | | |
| 90-day mRS 0-2 | 31 (35.6) | 47 (50.5) | 0.044 | 30 (33.3) | 48 (53.3) | 0.007 |
| Discharge NIHSS 0-1 | 17 (19.5) | 28 (30.1) | 0.102 | 15 (16.7) | 30 (33.3) | 0.010 |
| Composite | 33 (37.9) | 54 (58.1) | 0.007 | 32 (35.6) | 55 (61.1) | <0.001 |
